# Stakeholder-engagement on assessment of implementation considerations for food-policy interventions for prevention of overweight and obesity in Kenya and evaluation of the engagement process

**DOI:** 10.64898/2026.04.18.26351190

**Authors:** Mary Njeri Wanjau, Lucy Mecca, Rose Okoyo Opiyo, Sarah Mounsey, Kibachio Joseph Mwangi, J. Lennert Veerman, Lucy W. Kivuti-Bitok

## Abstract

**Background:** Increasing global prevalence of overweight and obesity underscores the need for context-specific evidence to guide preventive policy implementation. Previous modelling found that promoting healthy indigenous foods, implementing a 20% tax on sugar-sweetened beverages (SSBs), and introducing mandatory kilojoule menu labelling in formal-sector restaurants in Kenya were health-promoting, cost-saving, and cost-effective. We engaged stakeholders to assess additional implementation considerations relevant to decision-makers and to evaluate the stakeholder engagement process used in the modelling study.

**Methods:** Using the Assessing Cost⍰Effectiveness approach, we conducted a stakeholder-engaged study with national-level Kenya stakeholders recruited through purposive and snowball sampling. Through deliberative dialogue at a hybrid workshop, stakeholders assessed implementation considerations such as equity, feasibility and sustainability using a colour-coded scoring tool. We evaluated the engagement process using an anonymous survey covering seven stakeholder-engaged research domains. We analysed responses thematically.

**Findings:** Across the three interventions, most implementation considerations for feasibility, reach and impact, affordability, acceptability, and sustainability were assessed as medium or high. Industry acceptability of kilojoule labelling and SSB tax and affordability of kilojoule labelling to industry were rated low. Equity scores varied. Stakeholders proposed complementary measures that could raise ‘low’ ratings to favorable scores.

Clarity on stakeholder roles was identified as a key strength of the engagement process, while competing time commitments limited participation.

**Interpretation:** Stakeholder insights place prior economic evaluation findings within a broader policy relevant framework and provide context-specific evidence that could inform current fiscal/regulatory debates. Evaluation of the stakeholder engagement processes can strengthen public health research.

**Funding:** No funding

## Introduction

Globally, the growing burden of overweight and obesity continues to strain healthcare systems, particularly in low- and middle-income countries already facing persistent childhood malnutrition, infectious diseases and climate change. ^1-3^ In Kenya, approximately 47% of adult females and 29% adult males had overweight or obesity in 2021.^1^ Elimination of overweight and obesity in Kenya could increase the health-adjusted life expectancy by 2·3 (95% UI 2·0-2·8) years for females and 1·0 (95% UI 0·8-1·1) years for males.^4^ If the rise of overweight and obesity were halted by 2025 (Kenya’s reduction target), 6·8 million health-adjusted life years HALYs (135 HALYs per 1000 persons) and ∼US$3 billion healthcare costs could be saved (US$62 per capita) over the lifetime of the 2019 population, compared to recent trends continuing until 2044.^5^ By 2044, productivity gains resulting from a reduction in related mortality and morbidity was estimated at ∼US$5·8 billion (∼US$237 per capita).^5^

Changes in dietary patterns and nutrient intakes, fuelled by unhealthy food environments, contribute to the rising burden of overweight and obesity.^6-9^ Targeted and carefully designed policy interventions that aim to create healthy environments with low individual agency can influence entire populations and work to make healthier foods more accessible and affordable.^10-14^ Context-specific evidence of impact and cost effectiveness of interventions can help policy makers identify and prioritise interventions for the prevention and control of overweight and obesity.^15^

In previous work, we evaluated two stakeholder-identified broad strategies. The first was a ‘research-based strategy to increase consumption of healthy indigenous foods in Kenya’ which was modelled as two broad scenarios: (1) a reduction in national consumption levels scenario and (2) a change in consumption levels related to reduction in supermarket food purchase; the second was ‘creation of healthy food environment’ which was modelled as two specific food policies: (1) taxing sugar-sweetened beverages (SSBs) and (2) mandatory kilojoule menu labelling on food served in formal sector restaurants.^16^ We found that all interventions yielded substantial health gains, healthcare cost savings and productivity gains. Considering a health system perspective, the two specific interventions (a 20% SSB tax and mandatory kilojoule menu labelling) were assessed for cost-effectiveness and found to be dominant (health promoting and cost-saving). We proposed that these impactful and cost-effective interventions be considered for implementation in Kenya.^16^ In the current study, we engaged stakeholders with the aim of assessing other factors important to decision-makers in their considerations for the implementation of these interventions. The assessment places our evaluation results within a broader framework that seeks to incorporate other factors (implementation considerations) that are important to policy makers. Consistent with the Assessing Cost-Effectiveness (ACE) approach.^13,17,18^ Our findings may strengthen the policy relevance of our health and economic estimates for priority setting in overweight and obesity prevention in Kenya and similar settings. In addition, we conducted a survey to evaluate the stakeholder engagement process and information delivery in order to identify what worked well and what could be improved in future stakeholder-engaged modelling studies.

## Methods

### Stakeholder recruitment

Stakeholders were drawn from various institutions and sectors including the Kenya Ministry of Health, the Kenya National Research Institute, humanitarian aid, university and civil society organizations. Identification of stakeholders and recruitment process are reported elsewhere.^15,19^ In summary, we used purposive and snowball sampling techniques to recruit stakeholders involved policymaking at the national level. At the beginning of the stakeholder engaged modelling study, we prepared a description of potential stakeholders which guided our recruitment process (Supplementary File [SF] Table 1). Table 1 presents a summary and timeline of the stakeholder engagement conducted throughout the project.

**Table 1:**
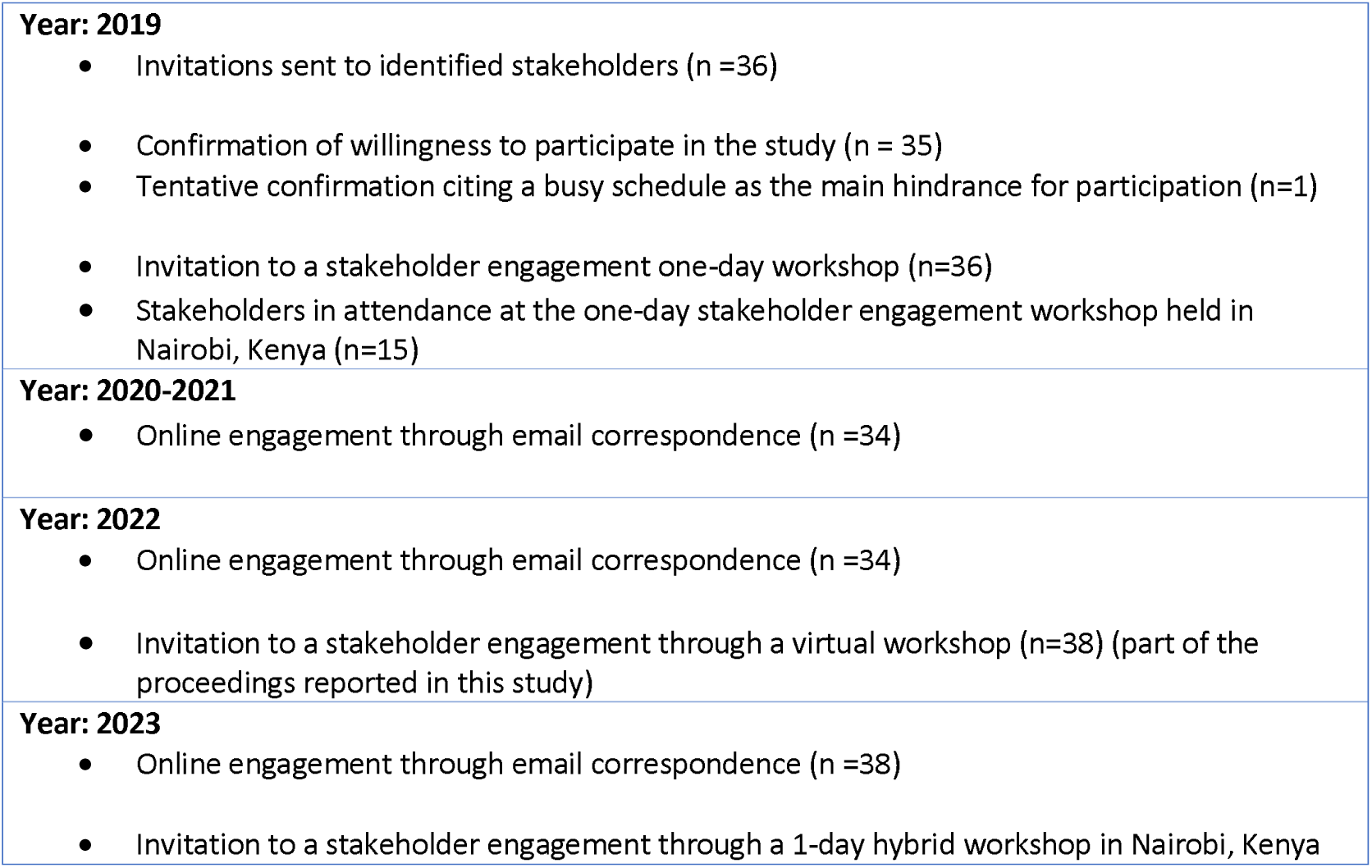
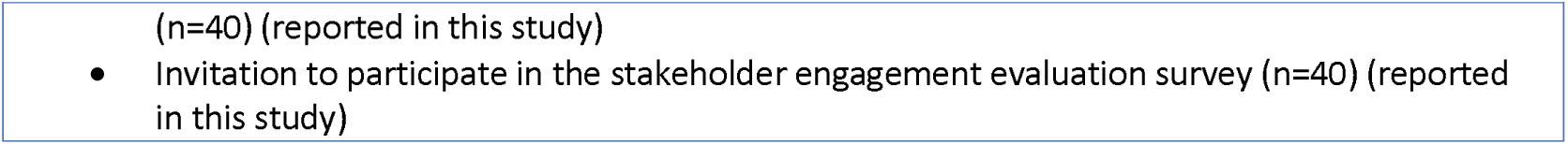
Summary and timeline of the stakeholder engagement.

| **Year: 2019**<br>• Invitations sent to identified stakeholders (n =36)<br>• Confirmation of willingness to participate in the study (n = 35)<br>• Tentative confirmation citing a busy schedule as the main hindrance for participation (n=1)<br>• Invitation to a stakeholder engagement one-day workshop (n=36)<br>• Stakeholders in attendance at the one-day stakeholder engagement workshop held in Nairobi, Kenya (n=15) |
|---|
| **Year: 2020-2021**<br>• Online engagement through email correspondence (n =34) |
| **Year: 2022**<br>• Online engagement through email correspondence (n =34)<br>• Invitation to a stakeholder engagement through a virtual workshop (n=38) (part of the proceedings reported in this study) |
| **Year: 2023**<br>• Online engagement through email correspondence (n =38)<br>• Invitation to a stakeholder engagement through a 1-day hybrid workshop in Nairobi, Kenya (n=40) (reported in this study)<br>• Invitation to participate in the stakeholder engagement evaluation survey (n=40) (reported in this study) |

### Research aim 1: Assessment of other implementation considerations for each food policy intervention evaluated in the larger study

In a 2·5-hour online meeting held in October 2022, the research team proposed to adapt some of the implementation considerations used in previous ACE studies for the assessment. ^13,17^ These considerations included equity / capacity to reduce health inequalities; feasibility of implementation; reach and size of impact; affordability; acceptability of intervention and sustainability (Table 2). The meeting was conducted and recorded on Microsoft Teams. Three authors (MNW, LV and LWK-B) facilitated the 45-minute through a deliberative dialogue process, an approach useful for managing potential over-representation of active participants and to ensure balanced participation.^15,19,20^ Stakeholders participated in value judgement aspects of the assessment of other factors or policy implementation considerations that are important to decision makers in Kenya. Key aspects of each consideration were reviewed and discussed for stakeholder input and buy-in. We used a traffic light colour coded scoring system for the assessment of each of the implementation considerations (Table 2).^13,17^ Each implementation consideration could be scored as being either high, medium or low for each intervention. The implementation consideration ‘equity’ would be scored as negative, neutral or positive. Following the online meeting, a copy of the assessment form was circulated to all stakeholders via email to obtain input from those who had not attended the meeting, as well as additional feedback from those who were in the meeting.

**Table 2:** Implementation considerations and categories for classifications.

| Implementation consideration | Key considerations<sup>#</sup> | Categories for classification |  |  |
|---|---|---|---|---|
| **Equity / capacity to reduce health inequalities** | Considers impact on the equity of distribution of disease and health status and access to, or utilisation of, specific interventions. | Positive | Neutral | Negative |
| **Feasibility of implementation** | Considers the likely feasibility of implementation for an intervention, based on local/national/international experience and/or parallel evidence. | High | Medium | Low |
| **Reach and size of impact** | Considerations based on the type and nature of intervention, the mechanism of intervention (e.g., mandatory regulations, voluntary regulations/guidelines); size of impact expected. | High | Medium | Low |
| **Affordability** | Considers affordability of intervention to the government | High | Medium | Low |
|  | Considers affordability of intervention to the industry | High | Medium | Low |
|  | Considers affordability of intervention to the general public | High | Medium | Low |
| **Acceptability of intervention** | Acceptability to government | High | Medium | Low |
|  | Acceptability to industry | High | Medium | Low |
|  | Acceptability to the general public | High | Medium | Low |
| **Sustainability** | Considers likely sustainability based on: the mechanism of intervention (e.g., mandatory regulations, voluntary regulations/ guidelines); the level of ongoing funding required; and the likelihood that the intervention will result in sustained behaviour change. | High | Medium | Low |
| **Other important considerations specific to each intervention such as other positive or negative impact that may result from intervention (apart from the health and economic impact included in the modelling)** |  |  |  |  |

<sup>#</sup> Definitions and key considerations are largely adapted from previous ACE studies.<sup>13,17,18</sup>

In June 2023, we held a one-day hybrid workshop in Nairobi, Kenya. In preparation for the workshop, the assessment form was recirculated to the stakeholders via email. Using the same deliberative dialogue process, stakeholders assessed each intervention against the listed factors or implementation considerations within a 2·5-hour session. Two authors (MNW and LWK-B) facilitated the deliberations. The agreed upon scores and deliberations were typed into the assessment form by one author (MNW) and projected on the screen for all to view as the deliberations progressed. The online engagement was conducted and recorded on Microsoft Teams. One author (MNW) listened to the workshop recording and checked that all deliberations on the assessment scores and comments given by the stakeholders had been recorded. Two additional authors (LWK-B and LV) had access to the raw data for review and checking.

### Research aim 2: Evaluation of the stakeholder engagement process and information delivery in the stakeholder engaged modelling study

We developed a survey tool guided by the exploratory framework for evaluating stakeholder engaged research proposed by the Ray and Miller ^21^ which outlines 6 areas of evaluation: 1) expectations from the engagement; 2) representation of stakeholders; 3) degree of involvement; 4) engagement channels and methods; 5) future expectations and 6) benefits and barriers to engagement. For the purposes of our study, we modified this framework and added a 7th area on information delivery in the stakeholder engaged modelling study as a whole. The tool was administered as an anonymous online survey using the Microsoft Office Forms. During the one-day hybrid, a printed version of the survey tool was distributed to stakeholders who preferred to complete the survey using paper and pen (with no names indicated). Stakeholders absent from the workshop were invited to complete the online survey tool within the next two working days after the hybrid workshop. To encourage responses, reminder emails were sent in the weeks following the workshop and the online form remained open for an additional six weeks. Responses were thematically analysed across the survey tool’s seven evaluation areas, compiled in MS Excel, and analysed manually in MS Word.. One author (MNW) led the data analysis; two additional authors (LWK-B and LV) reviewed the raw qualitative data sets and the thematic analysis for agreement. All data were de-identified. Data were stored in a university computer and backed up in Griffith University research data storage services. Written informed consent was obtained from each participant in accordance with the Declaration of Helsinki (https://www.wma.net/policies-post/wma-declaration-of-helsinki/).

Each participating stakeholder received Kshs. 3,000 (approx. USD25) via M-PESA to cover internet or transport costs for attending the online meeting or hybrid workshop. The study was approved by the Griffith University Human Research Ethics Committee (GU Ref No: 2019/707) and the Kenyatta National Hospital/University of Nairobi Ethics & Research Committee (P81/02/2021).

## Results

### Assessment of other implementation considerations

A total of 11 stakeholders attended the online meeting held on Microsoft Teams in October 2022 and 16 stakeholders (ten in person and six online) attended the June 2023 workshop. A list of stakeholders who participated by institution representation is provided in the SF Table 2. The assessment scores of other implementation considerations for each food policy intervention are presented in Table 3. In this section we also report on any stakeholder comments given for each of the scores. Other overall considerations given are reported in Table 3.

**Table 3:** Assessment scores for the implementation considerations for each intervention.

| **Intervention/ Implementation consideration** | **1. Research-based strategy to increase consumption of healthy indigenous foods** | **2. Mandatory kilojoule labelling on food served in formal sector restaurants** | **3. Tax on sugar-sweetened beverages (SSBs)** |
|---|---|---|---|
| **1.Equity / capacity to reduce health inequalities** | Neutral | Negative | Neutral |
| **2.Feasibility of implementation** | High | High (in formal sector) | High |
| **3.Reach and size of impact** | High | Medium | Medium |
| **4.Affordability of intervention** |  |  |  |
| **to the government** | High | High | High |
| **to the industry** | Medium | Low | Medium |
| **to the general public** | High | High | Medium |
| **5.Acceptability of intervention** |  |  |  |
| **to government** | High | High | Medium |
| **to industry** | High | Low | Low |
| **to the general public** | High | High | Medium Low |
| **6.Sustainability** | High | Medium Low | High |
| **Other considerations (verbatim quotes from participants)** |  |  |  |
| • Perhaps it is good to stratify modelling by socioeconomic classes or literacy levels as this may inform the impact of interventions. For example on whether people read and understand the kilojoule labelling.<br>• From the listed strategies and priorities [previously identified by stakeholders<sup>19</sup>], similar work linked to physical activity and transport related interventions is needed. A call for future research on these areas.<br>• Seek collaborations and funding for future research on how else the developed obesity model can be utilised.<br>• Stakeholders to play a role in results dissemination, distilling the policy briefs to help findings inform policy in Kenya. It was noted that all publications arising from this study are open access and can be read in full without requiring article purchase or journal subscription fees.<br>• Future research into the factors that influence in priority setting for non-communicable disease control in Kenya is needed. How can the factors be optimised or managed? [Factors identified by stakeholders<sup>15</sup>]. Development of policy framework that responds to the factors that influence NCD prioritising.<br>• Generate own data and evidence from Kenya. For example, disease cost data or database, evidence on intervention effect size in Kenya – from full intervention implementation or pilot studies. |  |  |  |

Definitions and key considerations are largely adapted from previous ACE studies.<sup>13,17,18</sup> Overall, this assessment was based on stakeholders’ input.

Equity scores varied. The research-based strategy to increase consumption of healthy indigenous foods was assessed as having a neutral impact on equity. The stakeholders commented that readily accessible healthy indigenous foods could reduce inequity. However, these foods could be expensive and some fast [processed] foods may be cheaper, especially in the informal sector in urban settings; the indigenous nature of foods and activity levels/ energy expenditure are community specific; some communities may have less access to the indigenous foods e.g., informal settlements or arid and semi-arid lands (ASAL) in Kenya impacted by climate change. The mandatory kilojoule labelling on food served in formal-sector restaurants was assessed as having a negative impact on equity because it was limited to formal sector, which is used predominantly by wealthier, urbanized sections of the population. They indicated that impact will be influenced by people’s attitudes towards reading kilojoule labels on meals, limited to those who can read and understand issues of calorie measures and dependent on what the labels looked like-would they be visual or text? Consideration that food establishments and merchants would pass the cost of calculating the caloric content of their menu options to consumers and thus distort the analysis was made. It was considered that pilot studies in Kenya would be needed. Stakeholders commented that the impact of equity could be more favorable if retailers responded by reducing the number of calories per meal and if health education was done alongside the labelling to increase awareness and urge the people to read the labels. The informal sector could be reached if mandatory labelling also targeted higher levels in the production or supply chain e.g., regulating the sausage manufacturers for the ‘smokies’ sold to vendors in the informal sector. However, it was noted that the cooking process plays a role too, e.g. reuse of cooking oil for fried foods in the informal sector. Taxing SSBs was assessed as having a neutral impact on equity. The stakeholders commented that it depended on the scope of the tax. For example, it is important to consider potential industry responses, such as if producers fully pass the resulting price increase to consumers or if they reformulate their products by reducing the sugar content to avoid the tax.

Feasibility of implementation for all three interventions was scored as high. For the research-based strategy to increase consumption of healthy indigenous foods, it was considered that a small study could be conducted as a starting point, followed by subsequent community-specific studies. For both the mandatory kilojoule labelling on food served in formal sector restaurants and the SSB tax, differences in literacy and awareness levels within communities in Kenya would impact feasibility.

The reach and size of impact for all three interventions was favorable. The research-based strategy to increase consumption of healthy indigenous foods scored a ‘high’ rating with comments that a pilot study could have small reach; the research should have representative samples; include participants from all communities, including those from urban and cosmopolitan areas; definition of indigenous foods to consider loss of diversity (species and varieties that have disappeared or been replaced) or increased diversity (where communities may have integrated a broader range of indigenous foods). Mandatory kilojoule labelling and SSB tax both scored a ‘medium’ rating with comments that reach and size of impact would be high for people who ate in the fast-food formal restaurants but low for those who did not and that the intake of SSBs may be more prevalent in urban areas.

Affordability to the government for all three interventions scored ‘high’. It was noted that although the research-based strategy was likely expensive and time consuming, the government, non-governmental organizations or other development partners/agencies could allocate funds to such a project. Affordability to the industry was favorable (scored a ‘medium’ rating) for the research-based strategy and SSB tax. It was noted that the industry could potentially experience high profits from sale of the healthy indigenous foods, positive changes across the whole food production chain such as increased marketing and industry may increase SSB prices to match the tax. Mandatory kilojoule labelling scored a ‘low’ rating on affordability to industry with a comment that if government provided measures that allowed industry to implement labelling at lower costs, this could be a ‘medium’ rating. Affordability to the public was scored ‘high’ for the research-based strategy since the research itself would not be a direct cost to the public. However, it was noted that the resulting healthy indigenous foods may be expensive. Mandatory kilojoule labelling scored a ‘high’ rating too, considering that it doesn’t cost the public any extra unless the industry pass on labelling costs to the consumers. SSB tax was scored a ‘medium’ rating.

Acceptability to government for all three interventions was assessed as favorable. The research-based strategy and mandatory kilojoule labelling scored ‘high’ and the SSB tax scored ‘medium’. Acceptability to industry was scored ‘high’ for the research-based strategy but ‘low’ for both mandatory kilojoule labelling and SSB tax. Comments were that acceptance may not be as high due to increased production costs in the estimation of the number of kilojoules in the food, preparation and presentation of labels. However, they noted that both mandatory kilojoule labelling and SSB tax are regulatory policies and industry compliance is anticipated. Acceptability to the public was scored ‘high’ for the research-based strategy as it was considered it would be appealing to the public and indigenous foods in upcountry regions are affordable. The mandatory kilojoule labelling was scored ‘high’ too where implementation was complemented with increased reach and/or improving health literacy around the intervention. Acceptability of SSB tax to the public was scored ‘medium’ and ‘low’ and no consensus was reached.

Sustainability of the intervention scored ‘high’ for the research-based strategy with a note that it could be ‘medium’ due to a generation gap. Younger population groups may not be familiar with the indigenous foods. The mandatory kilojoule labelling was scored ‘medium’ and ‘low’ with no consensus reached. It was considered a challenge in Kenya as credibility of the labels needed to be reinforced and sustained. Sustainability depended on the population uptake and belief in the labels-information provided needed to be trusted. SSB taxation received a ‘high’ rating for sustainability.

### Evaluation of the stakeholder engagement process and information delivery in the stakeholder engaged modelling study

Of the 40 stakeholders invited, nine completed the evaluation survey; three used the printed form at the workshop and six responded via the online form. Of the nine respondents, seven indicated that they were engaged since the first workshop consultation in 2019 while two joined at the one-day workshop held in 2023.

### Expectations from the engagement

All nine respondents indicated that their role as a stakeholder on this project had been clear. Their expectations of this study included adding value to the desired results and meeting the study objectives, learning more about the [study] findings, obtaining new knowledge on obesity and prevention in Kenya, [developing] an adaptable model, gain[ing] knowledge on what works best in the fight against obesity in Kenya and supporting research on lifestyle, behaviour and health. One participant said, “…*I came to the workshop to learn and interact with the best that the industry has to offer. I also, aspired to air my contributions towards the realisation of workshop objectives shared prior to the event.*”

Personal expectations that were met during their engagement related to their contribution to the study such as the identification of interventions to be evaluated in the study (n=3). One stakeholder described it as “*achieving consensus on priority interventions”.* Seven participants listed expectations related to learning, motivation and gaining new information from the study. Of the seven, responses included *“I got to learn a lot from a team of dedicated researchers in Kenya and Australia”*, *“from the modelling, new findings that could support future policy were realised*” and one described it as “*learning from others globally*”. Two listed networking and making new friends with similar interests and another said that “*stakeholders’ opinions were taken seriously*”.

Four participants indicated that they had no unmet expectations from their engagement in this study. The remaining five, stated that they had expected the inclusion of stakeholders living with overweight and obesity (n=2); research to get to the *mashinani (*grass root level); provision of county specific data on obesity parameters; have a wider pool of representative stakeholders and to have evaluated “*the impact of physical education (PE) in CBE* [Competency-Based Education policy in Kenya]*”* (this was one of the interventions identified in the 2019 workshop but ranked fourth^19^ and not modelled in our study due to limited time and resources^16^).

### Information delivery in the stakeholder engaged modelling study

All nine respondents considered the information delivered to them throughout the stakeholder engaged modelling study as trustworthy and credible. Quoted responses included *“…the discussions were open/frank. No coercion”, “credible and trustworthy. Evidence of work done and literature from previous research”, “the team did very well to find and detail data that is specific to the Kenyan set-up despite the obvious lack of documented information in certain areas* [example, some evidence on intervention effect size was from international sources^16^*]”* and one commended the research team on work published in *“several internationally acclaimed journals”*.

All nine respondents indicated that the information delivered in this study had been clear. One added that “*the results were especially very well presented*” and another said, “*It was well delivered and clarifications were made where we had questions.”*

All nine respondents said that the information/results of this study were helpful to them in their daily work and advocacy for NCD control in Kenya. One participant said *“…it is clear that obesity cannot be ignored and there are clear benefits for prevention”* and another *“…I believe that this study has added to the body of knowledge that will be useful when formulating policies at the national Ministry of Health in Kenya where I work.”*

### Representation of stakeholders

Six respondents indicated that there had been a good representation of stakeholders. Quoted responses from participants included *“a good representation from a wide spectrum of different professions/sectors”; “covers* [covered] *broad area of key institutions”; “fairly good. Some did not turn up”; “the number of stakeholders that turned up both physically and online was impressive. However, there is room to reach out to more stakeholders”;* “*I missed persons from KARLO* [Kenya Agricultural & Livestock Research Organisation*] and universities such as JKUAT* [Jomo Kenyatta University of Agriculture and Technology*] Applied Nutrition Program, UON* [University of Nairobi]*”; “broader representation; stratified according to location (urban/peri-urban and rural).”*

Participants listed the following types of stakeholders or sectors for inclusion in similar engagements in future: *members of parastatals* [state corporations], *president advisors, ministry of health, universities, researchers, county governors or CECs [*County Executive Committee*], people living with* [high] *BMI, “people struggling with overweight/obesity issues themselves (in case they were not intentionally targeted)”, Community Health Volunteers, NGOs [*Non-Governmental Organizations*], politicians, the education and agriculture sectors.* One participant had no comment on stakeholders for future inclusion and was satisfied that *“all stakeholders were well represented.”*

### Degree of involvement

All nine respondents indicated adequate engagement in the discussions. All participants were asked to suggest ways the research team could improve stakeholder involvement in the study. The responses included 1) *“consider people’s perceptions of food and indigenous food-values, beliefs and attitudes. Also consider meanings”*, 2) *“have a mid-term engagement during the research*” 3) “*development of original research question*” 4) *“perhaps scheduled feedback.”* Two participants said that the engagement had gone well and had been fully involved throughout the study process.

### Engagement channels and methods

All nine respondents expressed that engagement methods and channels utilized in this study had been appropriate. The combination of online and in person sessions was highlighted by two participants *“Besides the in-person and Zoom meetings, I was involved via email which worked very well for me. Good”* [online sessions had been on Microsoft Teams] and *“both virtual and face to face worked well.”* Six participants said that the number of interactions held, communications done (both online and face to face) and the duration of the engagement sessions had been adequate. Of the six, one added that one-hour online forums for dissemination/feedback/update engagements could be held regularly e.g. monthly. One participant said that *“The nature of the interactions were of high standards and the contributions from either side were significant. This being among the many interactions done and the quality of time that was provided for every session, I believe was optimum.”* However, two participants said that the number of interactions could have been more and another commented that online participants had been *“a bit quiet though they participated a bit.”*

### Benefits and barriers to engagement

All nine respondents said that the stakeholder engagement process was beneficial for NCD control in Kenya. Benefits cited included creating awareness through stakeholders, influencing NCD control policies and informing policy advocacy by relevant institutions. One respondent said *“I hope it will bring a true turnaround to the challenge of overweight/obesity* and another said that *“engaging people at policy level has the potential of catalysing change in policies.”*

It was also stated that the findings increased *“available tangible data [which] could go a long way in directing decision making processes and policies in Kenya”* and that the work paved way for future research, for example, *“. . . more in-depth analysis that covers specific regions and communities.”* One respondent said the process enhanced “*credibility. Stakeholder involvement is embedded in the Constitution, so the study is in compliance with this requirement.”*

Five participants said that they had not faced any challenges or barriers faced in the engagement process with one stating that “*the communications were prompt and on time and the recommendations provided* [by the stakeholders] *were duly noted* [by the research team]*.”* Of the remaining four participants, one cited *“online challenges*” and suggested “*availability of internet*” as a solution. Another participant mentioned “*academia/ research linkage is weak and silo mentality of institutions working on certain research topics”* and suggested *“engagement with all type of stakeholders including industry players. Collaboration/partners from more institutions”.* Another challenge given was *“competing interests over time”* and the participant suggested *“work with alternative meetings, invite extra people so the ‘remnants’ are reasonable in number*.” “*Non-attendance by some of the stakeholders”* was mentioned as a challenge and one participant suggested to “involve the stakeholders in workshop planning for date of meeting.” Of the five participants who identified no barriers, two responded to the survey question on solutions stating *“engagement. Some of us talk more than others”* and another said *“adequate notice for engagement. Feedback on the outputs from the engagement soon after the occurrence "*

### Future expectations

In Table 4, we present the participants thoughts and expectations on future engagement in research related to NCD control policies in Kenya.

**Table 4:**
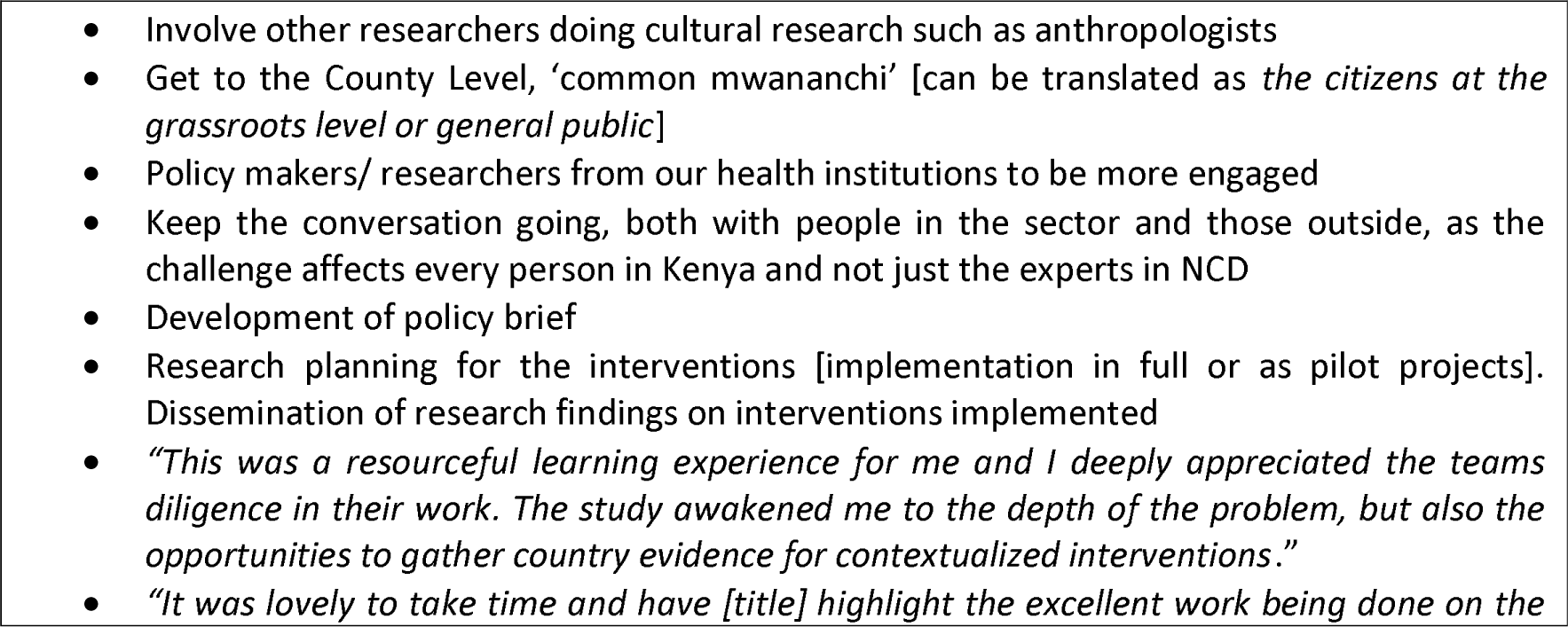
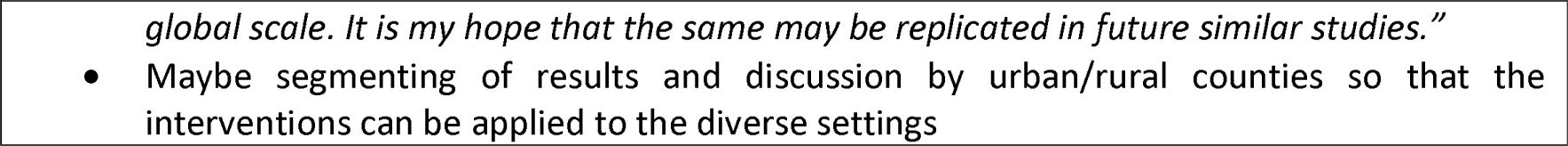
Thoughts and expectations of the stakeholders on future engagement in research or advocacy.

|  |
|---|
| • Involve other researchers doing cultural research such as anthropologists<br>• Get to the County Level, ‘common mwananchi’ [can be translated as *the citizens at the grassroots level or general public*]<br>• Policy makers/ researchers from our health institutions to be more engaged<br>• Keep the conversation going, both with people in the sector and those outside, as the challenge affects every person in Kenya and not just the experts in NCD<br>• Development of policy brief<br>• Research planning for the interventions [implementation in full or as pilot projects]. Dissemination of research findings on interventions implemented<br>• *“This was a resourceful learning experience for me and I deeply appreciated the teams diligence in their work. The study awakened me to the depth of the problem, but also the opportunities to gather country evidence for contextualized interventions*.”<br>• *“It was lovely to take time and have [title] highlight the excellent work being done on the global scale. It is my hope that the same may be replicated in future similar studies.”*<br>• Maybe segmenting of results and discussion by urban/rural counties so that the interventions can be applied to the diverse settings |

## Discussion

### Assessment of other implementation considerations

Our assessment of policy implementation considerations demonstrates stakeholder-engaged research (we aimed to answer research questions while engaging stakeholders as partners in our research, not as subjects of research).^16,19,21,23^ This was part of the due process in the Assessing Cost-Effectiveness (ACE) approach to priority setting^13,17,18^ applied in our larger study.^16^ The engagement allows for input beyond the research team and helps improve quality, transparency, relevance of study to policy and could accelerate adoption of findings into policy and practice.^15,21,24-29^ This study extends our previous evaluation results by integrating them within a broader framework that seeks to incorporate other factors (policy implementation considerations) important to decision-makers.

Implementation considerations on feasibility of implementation, reach and size of impact, affordability and acceptability to government, public and industry and sustainability were assessed as favorable (‘high’ or ‘medium’) for all interventions, except in five aspects: the acceptability of a 20% SSB tax to the public and sustainability of the mandatory kilojoule labelling were scored ‘medium’ / ‘low’ rating with no consensus reached towards either medium or low; affordability to industry for the mandatory kilojoule labelling and acceptability to industry for the mandatory kilojoule labelling and SSB tax were rated ‘low’. The research-based strategy to increase consumption of healthy indigenous foods and SSB taxation were both assessed as having a neutral impact on equity. The mandatory kilojoule labelling was assessed as having a negative impact because it is limited to the formal sector and relies on literacy, label design, and consumer engagement, but stakeholders noted that its equity impact could be improved through health education, product reformulation, piloting in Kenya, and extending regulation upstream in the supply chain to better reach the informal sector.

The research-based strategy to increase consumption of healthy indigenous foods was a broad, unique intervention and we did not find similar assessments of implementation considerations in the literature. As reported elsewhere, it was envisioned that research would be conducted to identify the nutritional value of indigenous foods, use the evidence to promote its production and ultimately increase consumption of healthy indigenous foods.^16,19^

The results of the assessment implementation considerations for mandatory kilojoule labelling and 20% tax on SSBs are comparable to results by Ananthapavan et al who found acceptability, feasibility and sustainability of both interventions in Australia favorable (‘high’ or ‘medium’) and acceptance of SSB tax by industry was scored a ‘low’ rating.^13^ Differences in scores were in the acceptability of the SSB tax to public, which Ananthapavan et al gave a ‘medium’ rating^13^ while we scored a ‘medium’ and ‘low’ rating. Like our study, the SSB tax was assessed as having a neutral impact on equity. ^13^ This is generally consistent with some international evidence that indicates stakeholders can sometimes have opposing ideas about the impact of SSB taxes particularly the economic impact on lower income groups (in the short term).^30-34^ While the government of Kenya progress towards implementing a health-based levy on SSB taxes, stakeholder views and insights into the challenges and facilitators of such a policy are valuable. Our previous estimates show that a 20% SSB tax in Kenya could cumulatively generate ∼71 billion USD in tax revenue by the 25^th^ year of implementation.^16^ If annual tax revenue were used to fund health initiatives in Kenya or to provide healthy food subsidies, the acceptability of the SSB tax to public and its impact on equity could be more favorable.

Ananthapavan et al scored mandatory kilojoule labelling a ‘medium’ rating for acceptability by industry and a ‘high’ rating for sustainability^13^ while we scored a ‘low’ rating for industry acceptability and a ‘medium’ and ‘low’ rating for sustainability. They ranked equity as neutral^13^ while we assessed it as having a negative impact, mainly due to the intervention targeting only formal restaurants. The differences in ratings could be due to the varying definition of the intervention in the two studies. Ananthapavan et al., evaluated the intervention as a mandatory legislation for fast food outlets to display energy content of foods and drinks on menus accompanied by a government sponsored education campaign.^13^ We evaluated mandatory kilojoule labelling on food served in formal sector restaurants. Kenya has a large informal food market and there could be different label literacy, different political economy and preferences to justify differences in acceptability/sustainability ratings between the two countries. In our study, stakeholders suggested that health education done alongside the labelling could enhance the impact on equity. They noted that high population uptake (reading of and acting on labels) and measures to reinforce and sustain the credibility of the labels could impact sustainability of the intervention. Other international studies on kilojoule/calorie labelling reported on menu labelling policies from upper-middle and high-income countries and did not include findings on policy implementation considerations.^35-37^

### Evaluation of the stakeholder engagement process

The evaluation of the engagement process demonstrates research on stakeholder engagement. We aimed to answer questions about engaging stakeholders^21^ i.e., identify what worked well and what could be improved in future stakeholder engaged modelling studies. The findings of this evaluation also reflect the strengths and limitations of our study.

All nine respondents indicated that their role as a stakeholder on this project had been clear. Their expectations of this study were aligned to the study objectives and roles of the stakeholders shared in the first workshop.^19^ We applied the deliberative dialogue approach and engaged facilitators to promote balanced participation. As indicated earlier, participants reported that they had been adequately involved in the discussions. We recruited stakeholders from the national policymaking level in line with our research aim since we evaluated policy interventions. ^19,24^ Future work focusing on the county level will provide insights into the prevention and control of overweight and obesity taking into account varying demographics and cultural aspects. We considered inclusion of stakeholders from civil societies to be a representation of the public. However, we acknowledge that inclusion of people living with overweight and obesity would add value though with a probability of stigma. Nonetheless, some of the stakeholders had lived experience with overweight, though that had not been part of our recruitment criteria. Most of the types of stakeholders suggested for inclusion in future had been included^19^ but had not turned up to the workshops and meetings due to multiple engagements and responsibilities. In a similar study conducted in Nepal, conflicting time commitments were identified as a major challenge to stakeholder engagement.^38^ In the initial workshop, stakeholders indicated that they preferred to hold workshops or meetings in the middle of the week with a very early morning start time. This advice was helpful in subsequent planning. In the evaluation, participants proposed additional measures to mitigate time limitations and non-attendance which could be implemented in future stakeholder engaged research. Suggestions to include community health volunteers, politicians, the education and agriculture sectors could be considered for inclusion in similar work in alignment to study aims.^24^

We experienced challenges with audio technology during the second workshop and those online could not hear the participants speaking in the room without the roaming microphone. Though online engagements were already included in the initial research plan and executed from 2019 (pre-Covid),^15,19^ three participants said that more engagements could have been held, suggesting, for example, holding regular one-hour online forums (e.g., monthly). Sharing of feedback on outputs soon after workshop and meetings was suggested. In the larger study, some of the workshop outputs had been shared inform of a report, in online meeting and emails and as manuscripts which had taken time to prepare. In future work, preliminary outputs could be shared with stakeholders promptly after an engagement. For example, this had been done after the 2022 online meeting where the assessment form for *other policy implementation considerations* was shared soon after the meeting. The stakeholders’ genuine enthusiasm in the study participation was commendable and inspiring. In a previous engagement, the stakeholders proposed establishing a joint communication platform, and the field researcher (MNW) was tasked with updating the ethics approval and setting up the platform.^19^ However, due to limited resources, the proposed platform was not set up. Similar to our study, other studies have also reported benefits in building connections amongst like-minded people and facilitating stakeholders’ advocacy for change and policy influence.^38-40^ As mentioned by one participant, Article 10 of the Constitution of Kenya (2010) firmly establishes inclusiveness, transparency, accountability and public participation (which includes stakeholder involvement), as core national values^41^ providing a constitutional foundation for stakeholder involvement in Kenya.

The study was impacted by the COVID 19 pandemic causing delayed approvals required from Kenya and cancellation of planned research travel to Kenya scheduled for 2020 and 2021. This study as a whole was not funded and costs (workshop venue costs, catering during the in-person workshop, stakeholders’ internet and transport facilitation fees, cost of purchase or printing of workshop materials) were supported by limited funds from a PhD project budget supplemented by principal supervisors’ (LV) research project funds. Despite these limitations, we established a network of stakeholders and maintained continued engagement throughout the study. Sustained stakeholder involvement throughout the research or implementation process has been identified as a positive outcome in other contexts.^38,39^ Stakeholders mentioned future expectations which included involvement of more groups such as anthropologists, the public, having more engagement from health institutions in similar studies, conducting further research at county level, implementation of the evaluated interventions in Kenya either in full or as pilot projects in the first instance and development of policy brief (s).

In Kenya, prevention and control of overweight and obesity is in its infancy with preventive food policies at the development stage.^42,43^ Our findings offer insights into policy implementation considerations for impactful, cost-effective food policy interventions that could help protect and create a healthy food environment in Kenya. The findings also contribute to enhancement of stakeholder engagement process in public health research. Benchmarking countries that have implemented similar food policies, such as South Africa’s SSB tax, could generate practical political-economy insights, inform strategies to manage industry responses that blunt the policy intentions, and guide on leveraging gains to strengthen health promotion, literacy, enforcement, and evaluation.

## Conclusion

Most of the policy implementation considerations for the interventions, research-based strategy to increase consumption of healthy indigenous foods in Kenya and creation of healthy food environment modelled through a 20% tax on sugar-sweetened beverages and mandatory kilojoule menu labelling on food served in formal sector restaurants, were assessed as favorable. Stakeholders suggested various complementary policy measures that could help raise ‘low’ rating to more favorable scores to facilitate possible implementation of these policies in Kenya. These interventions present potential solutions to overweight and obesity in Kenya. Benchmarking countries with similar food policies could inform context-specific design and implementation. Our study provides insights into the process of stakeholder engagement in public health research.

## Data Availability

The data supporting the findings of this study are available within the article and the Supplementary File

