## Supplementray File for "Stakeholder-engagement on assessment of implementation considerations for food-policy interventions for prevention of overweight and obesity in Kenya and evaluation of the engagement process"

**Methods**

**SF Table 1: Description of potential stakeholders**

Overall description

A team looking at preventive and early intervention strategies for NCD control; focusing on diseases or risk factors such as obesity. Or people involved in choosing the Public Health interventions to implement at any point in time- Priority setting either due to budget or any other considerations.

| Description of stakeholders |
| --- |
| • Head of Division NCDs, Ministry of Health (MoH), Kenya<br>• Head of Health Promotion Unit, MoH, Kenya<br>• Standards and Quality Assurance directorate, MoH<br>• A member (or members) from any health advisory committees recommended by the MoH Heads of divisions above.<br>• Other MoH officials - representatives from various divisions who would be involved in making choices of what interventions to implement and in what order. For example, officers from, health economics, data, and statistics.<br>• Representatives from other relevant agencies such as - Kenya Medical Research Institute (KEMRI), health economics body<br>• Representatives from influential and credible bodies that the MoH would recommend<br>• Representatives from Civil Society<br>• Medical Research Council representative<br>• An officer from the treasury who interacts with the health budget or activities<br>• A health counterpart in the Ministry of Planning, Ministry of Education, Science and Technology<br>• External partners for example, WHO health representative overseeing NCD control or health promotion<br>• Academic Experts in health systems management and health economics drawn from universities in Kenya |

### Results

#### SF Table 2: Stakeholders who participated in the 2022 online meeting and 2023 hybrid workshop by institution representation

**Representation of 11 stakeholders who attended the 2022 online meeting**

1. National Commission for Science, Technology, and Innovation
2. University of Nairobi, Kenya (n=2)
3. Strathmore University, Kenya
4. Personal consultant in Public Health - supply chain management
5. Kenya National Commission on Human Rights
6. Mater Hospital, Kenya
7. Kenyan Network of Cancer Organizations (n=2)
8. Ministry of Health (MoH), Kenya
9. Participant from a development agency attending in own capacity

**Representation of 16 stakeholders who attended the 2023 hybrid one day workshop**

**In person**

1. National Commission for Science, Technology, and Innovation
2. Institute of Diplomacy and International Studies, University of Nairobi, Kenya
3. Ministry of Health
4. Kenyan Network of Cancer Organizations (n=2)
5. International Institute of Legislative Affair (n=2)
6. School of Nursing Sciences, University of Nairobi, Kenya
7. Strathmore University, Kenya
8. Department of Anthropology, Gender and African Studies, University of Nairobi, Kenya

**Online**

9. Department of Community Health, University of Nairobi, Kenya
10. Personal consultant in Public Health - supply chain management
11. Participant from a development agency attending in own capacity
12. Ministry of Health
13. Public Health, Dental School, University of Nairobi, Kenya
14. Kenya National Commission on Human Rights
